# Early combination of sotrovimab with nirmatrelvir/ritonavir or remdesivir is associated with low rate of persisting SARS CoV-2 infection in immunocompromised outpatients with mild-to-moderate COVID-19: a prospective single center study

**DOI:** 10.1101/2024.07.15.24310384

**Authors:** I. Gentile, G. Viceconte, F. Cuccurullo, D. Pietroluongo, A. D’Agostino, M. Silvitelli, S. Mercinelli, R. Scotto, F. Grimaldi, S. Palmieri, A. Gravetti, F. Trastulli, M. Moccia, A.R. Buonomo

## Abstract

**Background:** Immunocompromised patients are at high risk of developing persisting/prolonged COVID-19 Data concerning early combined use of antivirals and monoclonal antibodies in this population are scarce.

**Research design and methods:** We performed an observational, prospective study, enrolling immunocompromised outpatient adults with mild-to-moderate COVID-19 treated with a combination of sotrovimab plus one antiviral (remdesivir or nirmatrelvir/ritonavir) within 7 days from symptoms’ onset.

**Results:** We enrolled 52 patients. No patient was hospitalized within 30 days from the disease onset, needed oxygen administration or died within 60 days, or experienced a reinfection or a clinical relapse within 90 days.

Clearance rates were 67% and 97% at 14th day after the end of therapy and at the end of follow-up, respectively.

Factors associated with longer infection were initiation of therapy after 3 days from symptoms onset, and enrollment more than 180 days from the beginning of the study. However, only the latter factor was independently associated with longer SARS-CoV-2 infection, suggesting a loss of efficacy of this strategy with the evolution of SARS-CoV-2 variants

**Conclusions:** Early administration of combination therapy with a direct antiviral and sotrovimab seems to be effective in preventing hospitalization, progression to severe COVID-19 and the development of prolonged/persisting SARS-CoV-2 infection in immunocompromised patients.

## 1. Background

COVID-19 still represents a major global health problem, especially for immunocompromised patients, in which a prolonged infection and greater risk of complications have been observed.[1] In particular, patients with impairment of humoral immunity (e.g., patients with B-cell hematologic malignancies or with a depletion of B-cells) manifest a protracted course of SARS-CoV-2 infection and shed viable virus for a longer period than immunocompetent patients.[1] Moreover, these patients have an increased risk of progressing to severe COVID-19 compared to the general population.[2] A prolonged or relapsing course not only results in increased attributable morbidity or mortality, but also delays chemotherapy and other therapeutic options such as stem cell transplantation, with negative impact on the outcome of the malignancy.[3]

Early treatment with antivirals or monoclonal antibodies prevents hospitalization and severe COVID-19 in fragile patients, but little is known about the role of such therapies in protecting highly immunosuppressed patients from developing prolonged/relapsing forms of COVID-19. Recently, some centers have described the use of a combination of monoclonal antibodies and one or two antivirals to treat COVID-19 in immunocompromised patients, but mostly in hospitalized patients, with severe COVID-19, and in patients that already developed persistent infection, with a predictable low impact on the disease’s outcome, once the persistent infection has already taken its course.[6–8]

Conversely, there is a scarcity of data in literature about the early use of combination therapy in early phase of COVID-19 in immunocompromised outpatients, for which an early administration of a combination therapy is expected to prevent the development of persistent COVID-19 and, therefore, the morbidity and mortality associated with it. Therefore, the aim of our study was to assess the efficacy of early combination treatment with one antiviral and a monoclonal antibody presumably active against the circulant SARS-CoV-2 variant, in non-hospitalized immunocompromised outpatients with mild-to-moderate COVID-19.

## 2. Patients and methods

### 2.1 Population

In an observational, prospective study, we consecutively enrolled immunocompromised adult patients, acceding the ambulatory service for COVID-19 outpatients of Federico II University Hospital in Naples, Southern Italy, from the 1st of May 2023 to 30^th^ December 2023.

### 2.1.1 Inclusion criteria

- Presence of at least one of the following active conditions:
  a. Primary immunodeficiency.
  b. Solid organ transplant on immunosuppressive therapy.
  c. chimeric antigen receptor T-cell therapy or allogeneic or autologous hematopoietic stem cell transplantation within 1 year.
  d. Acute myeloid/lymphoblastic leukemia within 6 months.
  e. Chronic lymphoblastic leukemia.
  f. Non-Hodgkin lymphoma within 1 year from last specific therapy.
  g. Plasma cellular neoplasms accompanied by hypogammaglobulinemia or receiving immunotherapy directed against B cells (bi-specific antibodies or antibody-drug conjugates against CD19, CD20 or BCMA).
  h. Primary or secondary hypogammaglobulinemia.
  i. Use of anti-CD20 for non-malignant conditions in the last 6 months.
- Mild-to-moderate COVID-19, with a SpO2≥94% on room air or on usual oxygen support, if already in use for chronic conditions.
- Symptoms onset ≤7 days.
- Diagnosis with SARS-CoV-2 nasopharyngeal swab (NFS) with real-time polymerase chain reaction (RT-PCR).
- Eligible to receive monoclonal antibody plus one antiviral between nirmatrelvir/ritonavir (N/r) or remdesivir.

### 2.1.2 Exclusion criteria

- SARS CoV-2 infection during the previous 3 months.
- Age younger than 18.
- Hospitalized patients.
- Incapable of giving written informed consent
- Patients with contraindications for antiviral.
- Patients who refuse to take any of the prescribed therapies.
- Patients who interrupt prematurely the treatment and
- Patients resulting SARS-CoV-2 negative during the treatment or within 3 days from the end of the therapy.

### 2.2 Outcomes

#### 2.2.1 Primary outcome

- Proportion of patients hospitalized for any reason within 30 days from the onset of the symptoms of COVID-19.

#### 2.2.2 Secondary outcomes

- Proportion of patients needing for oxygen administration or with an increase in oxygen flow if already on chronic oxygen therapy,
- Proportion of patients with prolonged viral shedding, defined as persistence of detectable SARS-CoV-2 on NFS below 34 Ct for 14 or more days from the end of the therapy.
- Death for any reason within 60 days from the onset of the symptoms of COVID-19.
- Reinfection or a clinical relapse within 90 days from the end of the therapy

### 2.3 Data collection

All the patients acceding to the outpatient service for COVID-19 were screened for eligibility. Eligible patients received signed informed consent and received a single intravenous dose of Sotrovimab 500 mg and either intravenous remdesivir (200 mg on day 1 and 100 mg i.v. on day 2 and 3) or oral N/r (300/100 mg twice daily for five days). The choice between the two antivirals was made by the prescribing physician based on patient’s history, estimated glomerular filtration rate (eGFR) and drug-drug interactions.

Patient who received remdesivir were scheduled for a second and a third dose on day 2 and 3, while patients eligible for N/r received the full tablet package to complete the therapy at home. In some cases, blood samples were requested on the first visit or during the follow-up to assess hepatic or renal function before or during therapy, or to measure inflammatory markers, whenever deemed appropriate for clinical reasons.

A mobile number available for 12h a day, 7 days a week, was provided to all the patient to report any issue regarding the therapy or the course of the disease. All the patients were followed up with weekly NFS for SARS-CoV-2 detection with RT-PCR until a negative result, meaning that SARS-CoV-2 was either not detectable or detectable with cycle threshold (Ct) above 34. Patients with Ct ≥ 34 and with no symptoms from at least 3 days were considered SARS-CoV-2 negative. Isolation precautions were interrupted in these cases and patients received formal clearance to continue immunosuppressant therapies, in case they were withheld for COVID-19 by the prescribers. Patients who decided to interrupt antiviral treatment prematurely, or that resulted SARS-CoV-2 negative during the treatment or within 3 days from the end of the therapy, were excluded from the study.

After reaching viral clearance, patients were instructed to inform the clinical center in case of any COVID-19 symptoms or in case of positive SARS-CoV-2 result on NFS performed for any reason. In these cases, patients received an appointment for clinical examination and repeated SARS-CoV-2 test with RT-PCR. In March 2024, every patient included during the observation period received a call from the investigators to specifically assess if they were still alive and if they had any reinfection or relapse. Data were crossed with the regional digital platform in which positive SARS-CoV-2 samples are recorded from general practitioners, hospitals, laboratories, and pharmacies.

### 2.4 Definitions

Prolonged SARS-CoV-2 infection was defined as persistence of detectable SARS-CoV-2 on NFS below 34 Ct for 14 or more days from the end of the therapy.

Monoclonal Antibody Screening Score (MASS) assigned a score to each of the original US FDA EUA criteria (released in November 2020) as follows: age ≥ 65 y (2), BMI ≥ 35 kg/m2 (2), diabetes mellitus (2), chronic kidney disease (3), cardiovascular disease in a patient 55 y and older (2), chronic respiratory disease in a patient 55 y and older (3), hypertension in a patient 55 y and older (1), and immunocompromised status (3). The maximum score is 18 [9,10].

### 2.5 Statistical analysis

Statistical analysis was performed using SPSS version 27 (SPSS Inc. Chicago, IL, USA). Continuous variables were reported as the median and interquartile range and categorical variables as the frequencies and percentages. Categorial variables were confronted with the chi-squared test and Fisher’s exact test, when appropriate. Continuous variables were confronted with the t-student test (parametric variables) or with the Mann-Whitney U test when nonparametric. A Kaplan-Meier analysis was performed to estimate the cumulative percentage of viral clearance over time among patients treated within 180 days and those treated later than 180 days from study onset. A Kaplan-Meier analysis was also performed to estimate the cumulative percentage of viral clearance over time among patients who started treatment for SARS-CoV-2 within 3 days from the onset of symptoms, and those treated later than 3 days from symptoms onset. Subsequently, the Cox Regression analysis was performed to calculate the risk factors for prolonged SARS-CoV-2 infection (> 14 days). All the covariates which resulted significantly associated with the dependent variable, or those with a p-value <0.2, were subsequently inserted in a multi-variate cox regression model to calculate the adjusted hazard ratio (aHR). The confidence interval was set at 95% for the interpretation of the results. A significance level of 0.05 was set for the interpretation of the results.

## 3. Results

During the study period, a total of 253 patients were screened for inclusion criteria, and 52 were considered eligible, then included and received combination treatment. No patients decided to interrupt prematurely the treatment and no patients resulted SARS-CoV-2 negative during the treatment or within 3 days from the end of the therapy.

As displayed in **Table 1**, patients had a median age of 63 years, were mostly vaccinated for SARS-CoV-2 (92%) with a median of 3 doses, and a low prevalence of comorbidities except for immunosuppressive conditions (median MASS 5/18), among which the most represented were hematologic malignancies (67%).

**Table 1.**
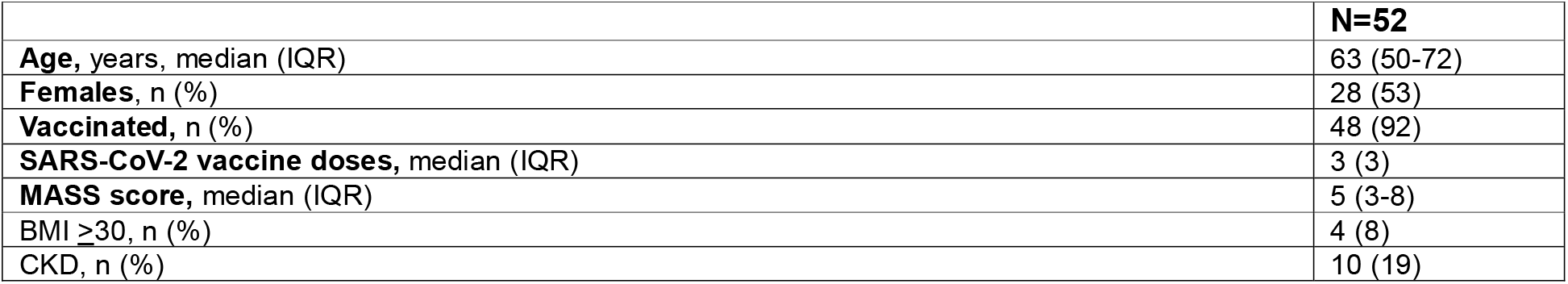
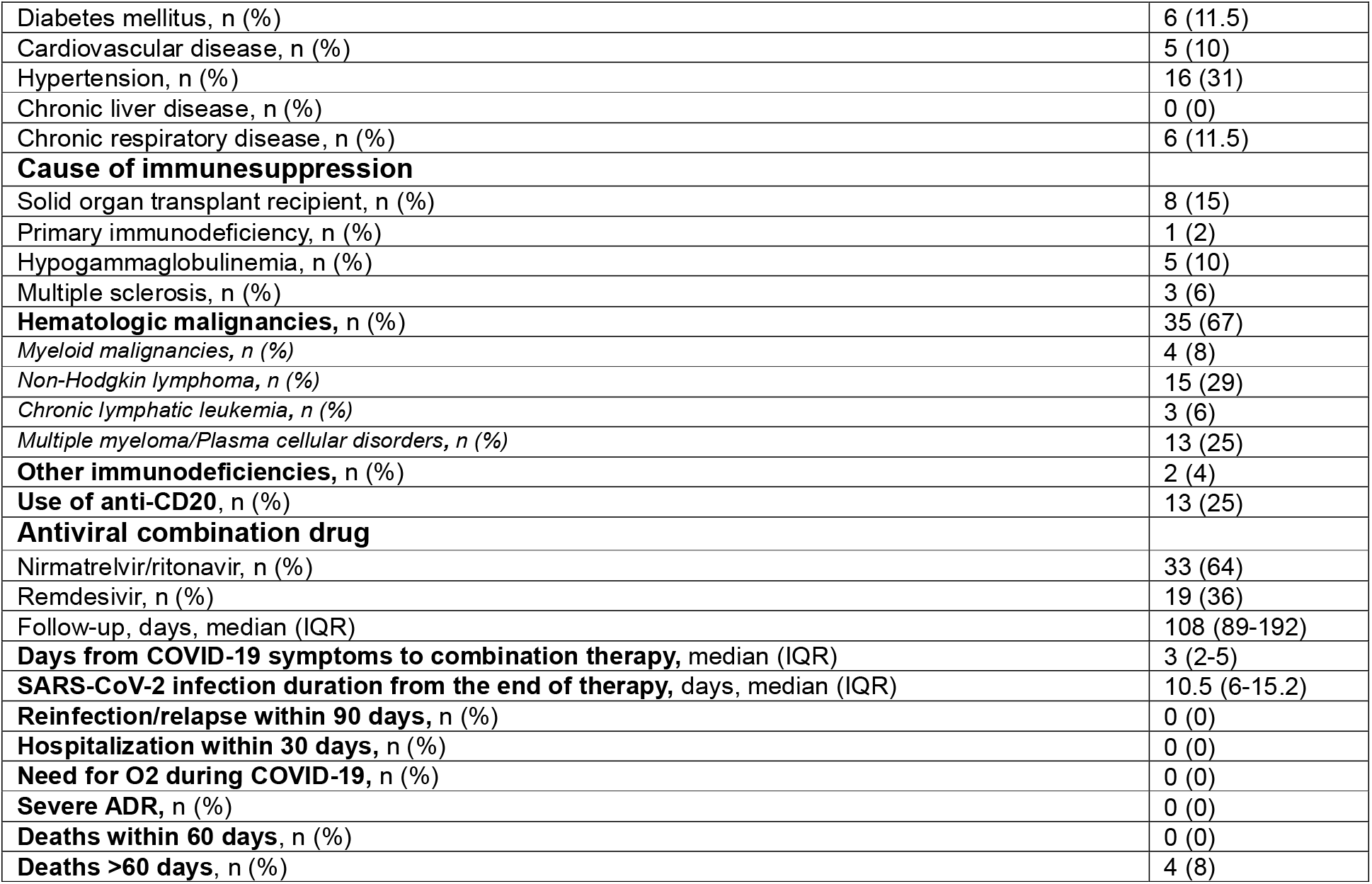
Demographic and clinical characteristics. MASS: Monoclonal Antibody Screening Score; BMI: body mass index; CKD: chronic kidney disease; ADR: adverse drug reaction.

After a median of 3 days (IQR 2-5) from COVID-19 symptoms onset, patient started combination treatment (N/r in 64% of cases and remdesivir in 36%). No patient was hospitalized within 30 days from symptoms’ onset, needed for O2 administration at home, died within 60 days or experienced a reinfection or a clinical relapse within 90 days. Four patients died 60 days after the beginning of the therapy, for causes unrelated to COVID-19, of which three were negative for SARS-CoV-2 at NSF, and one died with unknown SARS-CoV-2 status, since he unattended the follow-up after 30 days from therapy administration.

The median duration of SARS-CoV-2 infection from the end of the combination therapy was 10.5 days (IQR 6-15.2) and 17/53 (33%) of the patients had a prolonged infection (≥14 days from the end of the therapy). Nonetheless, nearly all the patients (50/51, 97%) reached documented viral clearance at the end of follow-up, except the abovementioned patient who died 58 days after therapy with unknown SARS-CoV-2 status and another patient that had detectable SARS-CoV-2 at the end of follow-up.

As shown in **Table 2**, patients with prolonged infection were not significantly different with respect to age, sex, vaccination coverage, type of underlying conditions, MASS score and type of combination therapy. Notably, prolonged infection occurred in 11/17 (64%) of patients who received antiviral treatment more than three days from symptoms’ onset, compared to 12/35 (34%) of patients who received treatment within three days (p<0.05). Moreover, we found no differences in terms of viral clearance between the use of nirmatrelvir/ritonavir and the use of remdesivir. We observed no severe adverse drug reactions and no patient stopped treatment because of adverse effects.

**Table 2.**
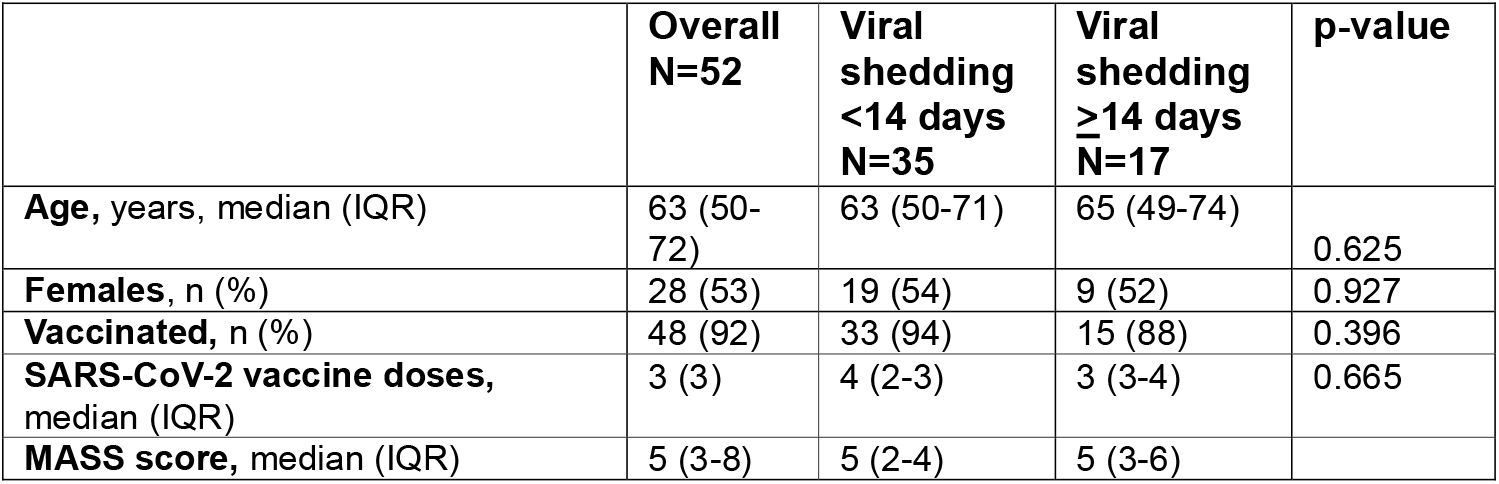
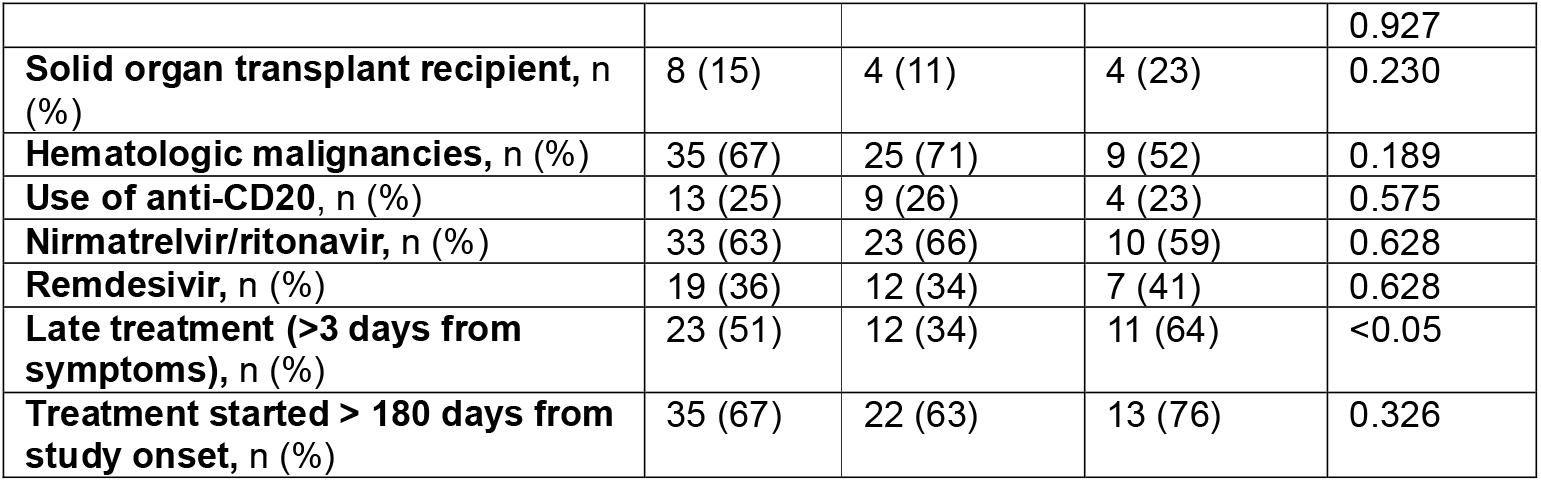
Factors associated with viral shedding longer than 14 days. MASS: Monoclonal Antibody Screening Score.

|  | **Overall**<br>**N=52** | **Viral**<br>**shedding**<br>**<14 days**<br>**N=35** | **Viral**<br>**shedding**<br>**≥14 days**<br>**N=17** | **p-value** |
| --- | --- | --- | --- | --- |
| **Age**, years, median (IQR) | 63 (50-72) | 63 (50-71) | 65 (49-74) | 0.625 |
| **Females**, n (%) | 28 (53) | 19 (54) | 9 (52) | 0.927 |
| **Vaccinated**, n (%) | 48 (92) | 33 (94) | 15 (88) | 0.396 |
| **SARS-CoV-2 vaccine doses**, median (IQR) | 3 (3) | 4 (2-3) | 3 (3-4) | 0.665 |
| **MASS score**, median (IQR) | 5 (3-8) | 5 (2-4) | 5 (3-6) |  |
|  |  |  |  | 0.927 |
| **Solid organ transplant recipient, n (%)** | 8 (15) | 4 (11) | 4 (23) | 0.230 |
| **Hematologic malignancies, n (%)** | 35 (67) | 25 (71) | 9 (52) | 0.189 |
| **Use of anti-CD20, n (%)** | 13 (25) | 9 (26) | 4 (23) | 0.575 |
| **Nirmatrelvir/ritonavir, n (%)** | 33 (63) | 23 (66) | 10 (59) | 0.628 |
| **Remdesivir, n (%)** | 19 (36) | 12 (34) | 7 (41) | 0.628 |
| **Late treatment (>3 days from symptoms), n (%)** | 23 (51) | 12 (34) | 11 (64) | <0.05 |
| **Treatment started > 180 days from study onset, n (%)** | 35 (67) | 22 (63) | 13 (76) | 0.326 |

Furthermore, we observed a notable variation in the duration of SARS-CoV-2 infection based on the enrolment time from the commencement of the study, as well as based on time of treatment initiation from the onset of symptoms. Specifically, patients who were enrolled more than 180 days after the study began (corresponding to the month of October 2023), exhibited significantly longer SARS CoV-2 infection (Log-rank 8.908, p<0.01) (**Table 3 Figure 1)**. Furthermore, patients who received treatment for SARS-CoV-2 infection later than 3 days from the onset of symptoms, also exhibited significantly longer SARS-CoV-2 infection (Log-rang 5.095, p<0.05) (**Table 4. Figure 2**). At the multivariate Cox regression analysis, enrollment of patients more than 180 days from the beginning of the study was the only variable independently associated with longer SARS-CoV-2 infection (aHR: 1.94; 94%CI: 1.05-3.56, p<0.05) (**Table 5**).

**Table 3.** Estimated time to SARS-CoV-2 viral clearance depending on time from study onset.

| time from study onset (days) | Median |  |  |  |
| --- | --- | --- | --- | --- |
|  | Estimate | Standard Error | 95% Confidence Interval |  |
|  |  |  | Lower Limit | Upper Limit |
| **≤ 180 days** | 6.000 | 2.236 | 1.617 | 10.383 |
| **> 180 days** | 14.000 | 1.389 | 11.277 | 16.732 |
| **Overall** | 11.000 | 1.078 | 8.886 | 13.114 |

**Table 4.** Estimated time for SARS-CoV-2 viral clearance based on the timing of treatment initiation from the onset of symptoms.

| time from study onset<br>(days) | Median |  |  |  |
| --- | --- | --- | --- | --- |
|  | Estimate | Standard Error | 95% Confidence Interval |  |
|  |  |  | Lower Limit | Upper Limit |
| ≤ 3 days | 11.000 | 1.326 | 8.401 | 13.599 |
| > 3 days | 14.000 | 1.797 | 10.478 | 17.522 |
| Overall | 11.000 | 1.078 | 8.886 | 13.114 |

**Table 5.** Cox regression analysis on risk factors for achieving viral clearance later. MASS: Monoclonal Antibody Screening Score.

|  | Univariate Analysis |  |  | Multivariate analysis |  |  |
| --- | --- | --- | --- | --- | --- | --- |
|  | HR | 95%CI | p-value | aHR | 95%CI | p-value |
| **Age**, years, median (IQR) | 0.99 | 0.95-1.03 | 0.709 | - | - | - |
| **Male sex**, n (%) | 0.77 | 0.29-2.01 | 0.619 | - | - | - |
| **Vaccinated**, n (%) | 0.37 | 0.07-1.81 | 0.220 | - | - | - |
| **SARS-CoV-2 vaccine doses**, median (IQR) | 1.60 | 0.43-5.91 | 0.483 | - | - | - |
| **MASS score**, median (IQR) | 1.04 | 0.87-1.26 | 0.631 | - | - | - |
| **Solid organ transplant recipient**, n (%) | 3.53 | 0.97-12.88 | 0.056 | 1.063 | 0.43-2.60 | 0.893 |
| **Hematologic malignancies**, n (%) | 0.39 | 0.13-1.16 | 0.091 | 0.93 | 0.47-1.84 | 0.853 |
| **Use of anti-CD20**, n (%) | 1.00 | 0.30-3.28 | 0.998 |  |  |  |
| **Nirmatrelvir/ritonavir**, n (%) | 0.55 | 0.20-1.53 | 0.258 | - | - | - |
| Remdesivir, n (%) | 1.80 | 0.65-4.94 | 0.258 | - | - | - |
| Treatment started > 180 days from study onset, n (%) | 2.10 | 1.15-3.83 | <0.05 | 1.94 | 1.05-3.56 | <0.05 |
| Late treatment (>3 days from symptoms), n (%) | 1.96 | 1.06-3.63 | <0.05 | 1.85 | 0.99-3.45 | 0.054 |

**Figure 1.**
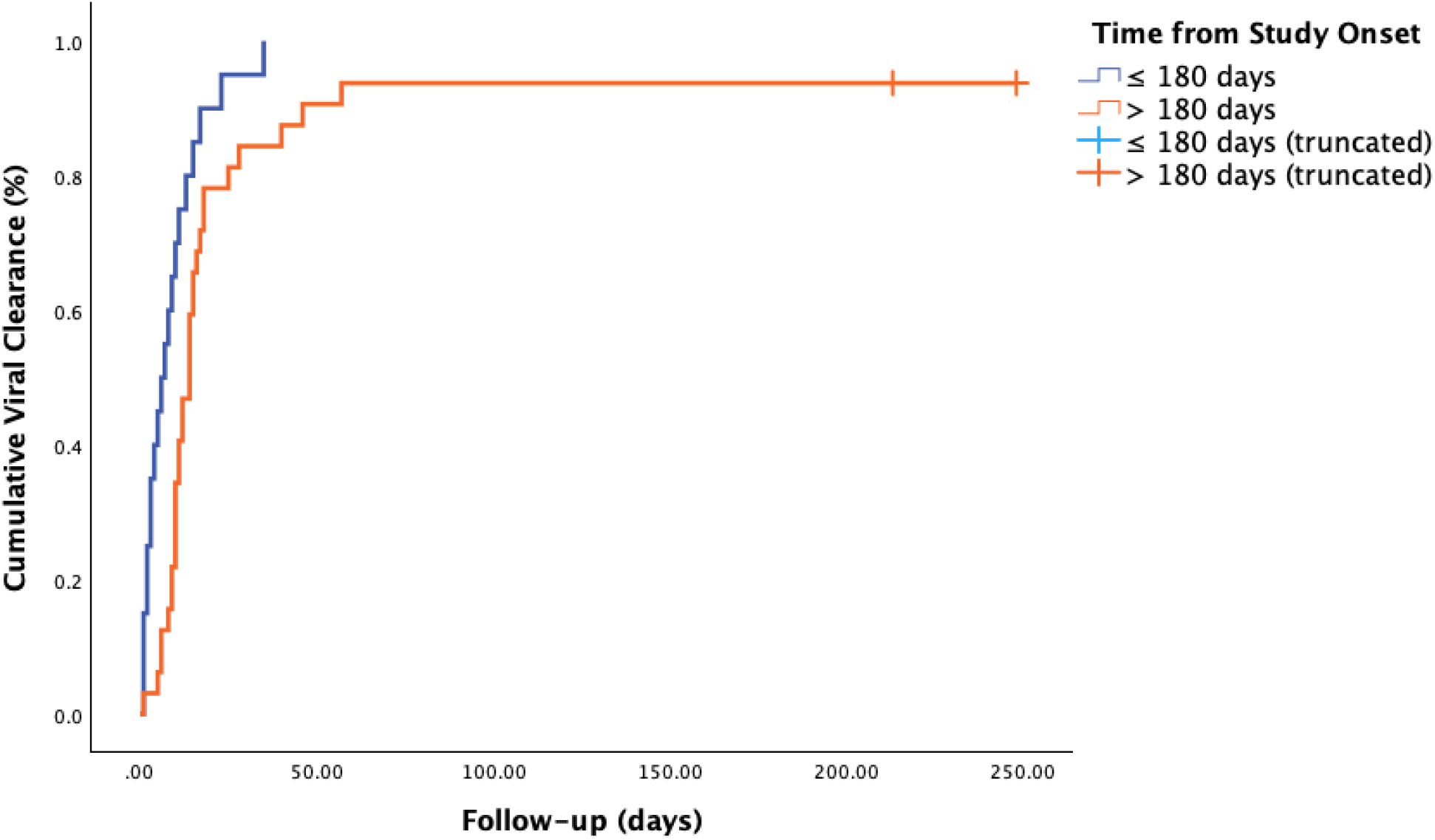
Kaplan-Meier analysis according to time of enrolment from study onset. Log-rank 8,908, p<0.01

**Figure 2.**
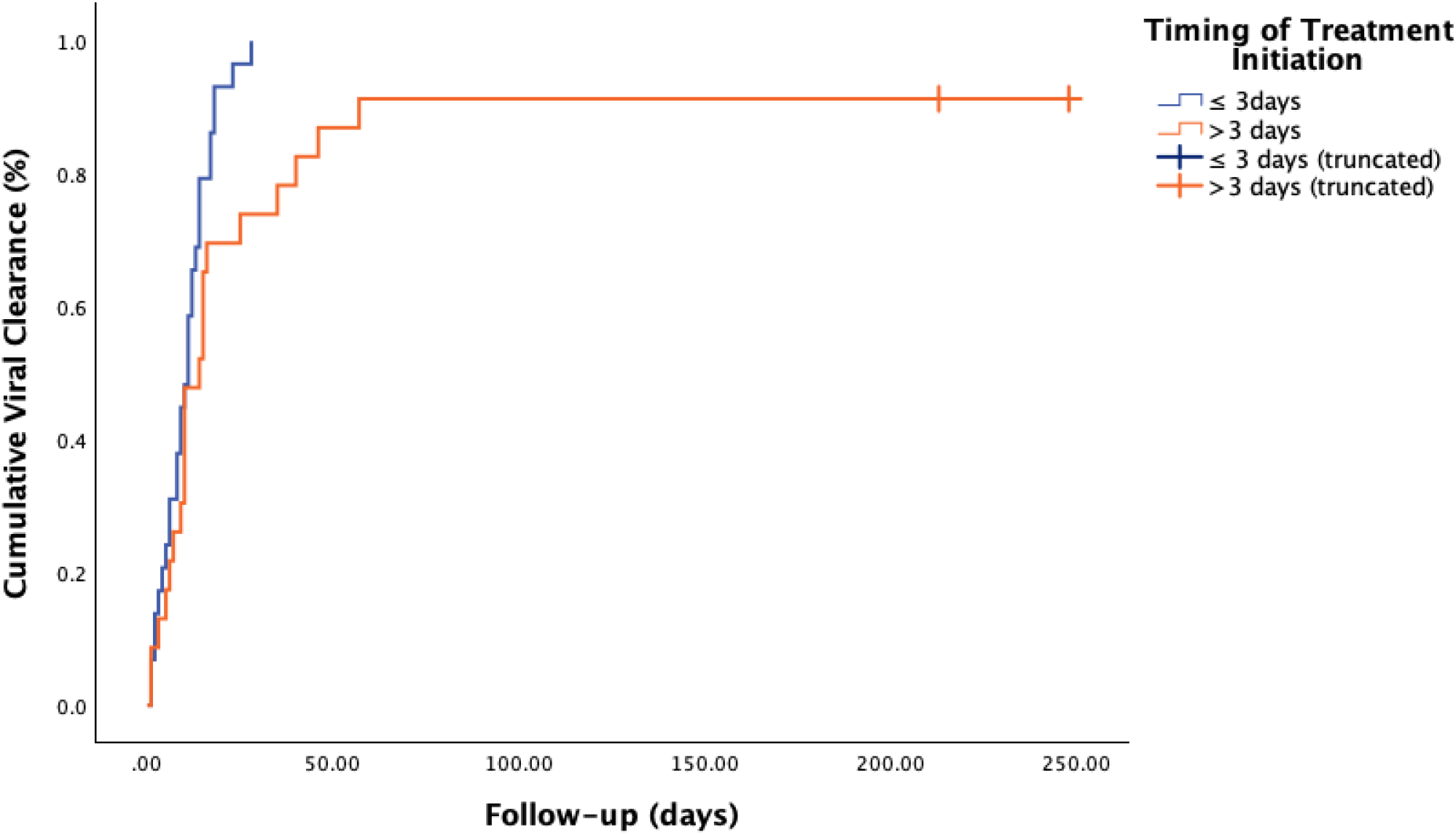
Kaplan-Meier analysis according to timing of treatment initiation from symptoms onset. Log-rank 5.095, p<0.05

## 4. Discussion

According to our results, early administration in the outpatient setting of combination therapy with one direct antiviral agent and the monoclonal antibody sotrovimab is associated with high viral clearance, low risk of death and of hospitalization, in a cohort of immunocompromised patients with mild-to-moderate COVID-19. In fact, none of our patients required hospital admission, oxygen therapy or died within 60 days from therapy administration for causes related to COVID-19. Moreover, the median duration of SARS-CoV-2 infection in our cohort was 10.5 days (IQR 6-15.2) and only 32% of the patients had a viral shedding longer than 14 days from the end of the therapy, with 96% of patients reaching stable virological clearance, with no relapse or reinfections during follow-up.

To date, only few authors have studied the systematic use of combination therapies in immunocompromised subjects, and these studies mostly reported the use of combination in patients that had already developed prolonged or persistent COVID-19, mostly hospitalized, for which the therapeutic effect of virus targeted therapies was already expected to be low [3].

For example, Mikulska and colleagues reported the use of combination therapy only in hospitalized patients, after a median time of 42 (IQR 29–100) days from SARS-CoV-2 infection, with a response rate at day 14, day 30, and last follow-up of 75%, 73%, and 82%, respectively [6]. Similarly, D’Abramo and colleagues have recently reported the use of combination therapy in a cohort of 69 immunosuppressed patients, hospitalized for severe COVID-19 (92% required oxygen therapy) and treated a median of 21 (IQR 8–36) days from symptoms’ onset [11]. Interestingly, in this study, the use of monoclonal antibodies (tixagevimab/cilgavimab or sotrovimab) in the antiviral combination was associated with a significantly higher rate of viral clearance. [11]. In both the abovementioned studies, the duration of viral shedding was longer than in our study, but treatment was started later during the course of infection.

On the other hand, a recently published paper by our group analyzed the efficacy and safety of the combination of 2 antivirals, with or without a mAb, both in early (within 10 days from symptoms) and in later phase (after 10 days) of SARS-CoV-2 infection in immunocompromised subject, finding that 100% of the patients treated early reached virological clearance at day 30 from the end of the therapy and were alive and well at follow-up, whereas corresponding figures in the late treated patients were 50% and 75%, with patients in late group more frequently needing for oxygen supplementation (p=0.015), steroid therapy (p=0.045) and reaching higher COVID-19 severity (p=0.017) [8].

In line with this, Orth and colleagues have recently presented the largest cohort (144 subject, of which 82% immunocompromised) of patients treated with combination therapy, so far [7]. According to co-primary endpoints (prolonged viral shedding at day 21 after treatment initiation and days with SARS-CoV-2 viral load□≥□10^6^ copies/ml), the authors found that underlying hematological malignancies and treatment initiation later than five days after diagnosis were significantly associated with longer viral shedding [7]. This finding has been confirmed and consolidated by our results, since we found a significant higher proportion of patient with prolonged infection (64%) among those who started antiviral therapy later than 3 days from symptoms (p<0.05). The delay in administering the combination therapy was significantly associated with a longer SARS-CoV-2 infection (HR 1.96; 95%CI 1.06-3.63, p<0.05), although this result was not confirmed at multivariate analysis.

Interestingly, we observed that the independent risk factor for not achieving SARS-CoV-2 clearance in our study was the enrollment in the last months of the study (from October to the end in December. We speculate that this could be related to a loss of efficacy of sotrovimab against the new circulating variants of SARS-CoV-2. In fact, during the study time, there has been a shift from a prevalent circulation of XBB1.5 variant, against which sotrovimab retained in vitro efficacy, towards new variants (XB1.9, BA2.86 and JN1), against which sotrovimab has shown higher 50% inhibitory concentrations (IC50) and therefore potential lower neutralizing activity [12,13]. Such consideration is of course underpowered by the fact that we didn’t assess sequenced the SARS-CoV-2 strains by which the patients were infected, and we didn’t test the neutralization capacity of sotrovimab in each case.

Another important limitation of the study is the lack of a control group of patients with similar immunosuppression, but treated with a single therapy, to assess the real advantage of a combination of antiviral and sotrovimab, compared to the antiviral alone.

Finally, we underline that in our study we used two different direct antivirals with different mechanisms of action together with sotrovimab. However, we did not find any difference in terms of main outcomes between the 2 different antivirals employed.

## 5. Conclusions

Despite its limitations, our study suggests that early administration of combination therapy with sotrovimab and a direct antiviral agent is safe and could be effective in preventing hospitalization, progression to severe COVID-19 and development of prolonged/persisting SARS-CoV-2 infection in severely immunocompromised patients. Circulation of new variants could prevent the efficacy of this strategy due to loss of efficacy of sotrovimab. Further studies are required to compare the combination approach with monotherapy in these categories, especially in light of the reduced activity of the monoclonal compound.

## Data Availability

All data produced in the present study are available upon reasonable request to the authors

## Author’s Contributions

IG, ARB and GV conceptualization; RS methodology and software; FC, DP, ADA, MS, SM, FG, SP, AG, FT, MM resources, formal analysis, investigation; GV, ARB, RS writing original draft; FT, IG, MM Writing - Review & Editing

## Ethics approval and consent to participate

All procedures performed in studies involving human participants were in accordance with the ethical standards of the institutional and/or national research committee and with the 1964 Helsinki declaration and its later amendments or comparable ethical standards. The study was approved by the Ethical Committee of the University of Naples Federico II (protocol n. 98/22).

## Consent for publication

written patient’s consent was obtained prior the enrollment to the study The data are recorded by the investigators in an anonymous manner such that subjects cannot be identified directly or through identifiers linked to the subject.

## Availability of data and materials

The data that support the findings of this study are available from the corresponding author upon reasonable request.

